# Retrospective screening for SARS-CoV-2 among 5,800 hospitalizations related to influenza-like illness during the 2018-19 pre-pandemic and 2019-2020 pandemic influenza seasons in the VAHNSI network, Spain

**DOI:** 10.1101/2021.05.24.21257402

**Authors:** Ainara Mira-Iglesias, Beatriz Mengual-Chuliá, Laura Cano, Javier García-Rubio, Miguel Tortajada-Girbés, Mario Carballido-Fernández, Juan Mollar-Maseres, Germán Schwarz-Chavarri, Sandra García-Esteban, Joan Puig-Barberà, Javier Díez-Domingo, F. Xavier López-Labrador, for the Valencia Hospital Network for the Study of Influenza and Respiratory Viruses Disease

## Abstract

On March 9 2020 the WHO Global Influenza Program (GIP) asked participant sites on the Global Influenza Hospital Surveillance Network (GIHSN) to contribute to data collection concerning severe acute respiratory syndrome coronavirus 2 (SARS-CoV-2). We re-analysed 5,833 viral RNA archived samples collected prospectively from hospital admissions for influenza-like illness (ILI) in the Valencia Region of Spain by the VAHNSI network (4 hospitals, catchment area population 1,118,732) during the prepandemic 2018/2019 (n=4,010) and pandemic 2019/2020 (n=1,823) influenza seasons, for the presence of SARS-CoV-2. We did not find evidence for community-acquired SARS-CoV-2 infection in hospital admissions for ILI in our region before early March 2020.

## Introduction

The first case of coronavirus disease 2019 (COVID-19) due to severe acute respiratory syndrome coronavirus (SARS-CoV-2) was detected in Wuhan (Hubei province, China) at the end of 2019 and the virus rapidly spread to other Chinese provinces and countries (1). As of 3 June 2020, the World Health Organization (WHO) had already reported the presence of SARS-CoV-2 in 54 different European countries (2). In Spain, the first imported and locally acquired COVID-19 cases were detected on 31 January, and 26 February 2020, respectively (3, 4). Since then, until beginning of June, Spain accounted for 240,660 confirmed cases and 27,133 deaths. Among total registered deaths, 86% were in people aged 70 or older and 95% had previous underlying chronic conditions (5). Symptomatology described by SARS-CoV-2-infected patients was almost identical to the common influenza-like illness (ILI) symptoms (6, 7), and COVID-19 cases may have occurred in Spain earlier than previously recognized (8). Besides, there is no data available on potential SARS-CoV-2 circulation during the prepandemic 2018/2019 influenza season.

## Methods

Every influenza season, from November to March/April, the Valencia Hospital Surveillance Network for the Study of Influenza and Other Respiratory Viruses (VAHNSI) network conducts a prospective active-surveillance study on respiratory viral infections in 4 hospitals, providing healthcare to 1,118,732 (22% of total) inhabitants from the Valencia Region of Spain (Figure 1). Our network is integrated in the Global Influenza Hospital Surveillance Network (GIHSN) sharing data with the WHO GIP (www.gihsn.org/). Full-time dedicated nurses screen all hospitalized patients discharged from the Emergency Department if they referred a complain possibly related to a respiratory infection. Patients are included in the study if they are resident in the catchment area of one of the participating hospitals, non-institutionalised and not discharged from a previous admission in the last 30 days. Patients have to meet the ECDC influenza-like illness (ILI) case definition (9), defined as the presence of, at least, one systemic symptom (fever or feverishness, malaise, myalgia or headache) and, at least, one respiratory symptom (shortness of breath, sore throat or cough), with an onset of symptoms in the 7 days prior to admission. For children less than 5 years old, we only require an onset of one symptom (Table S1) in the 7 days prior to admission. Finally, patients need to be in hospital between 8 and 48 hours to be enrolled and swabbed (combined nasopharyngeal and oropharyngeal in universal transport medium), after written informed consent. Clinical and demographic characteristics from patients were obtained by a face-to-face interview or by consulting medical records.

**Figure 1.**
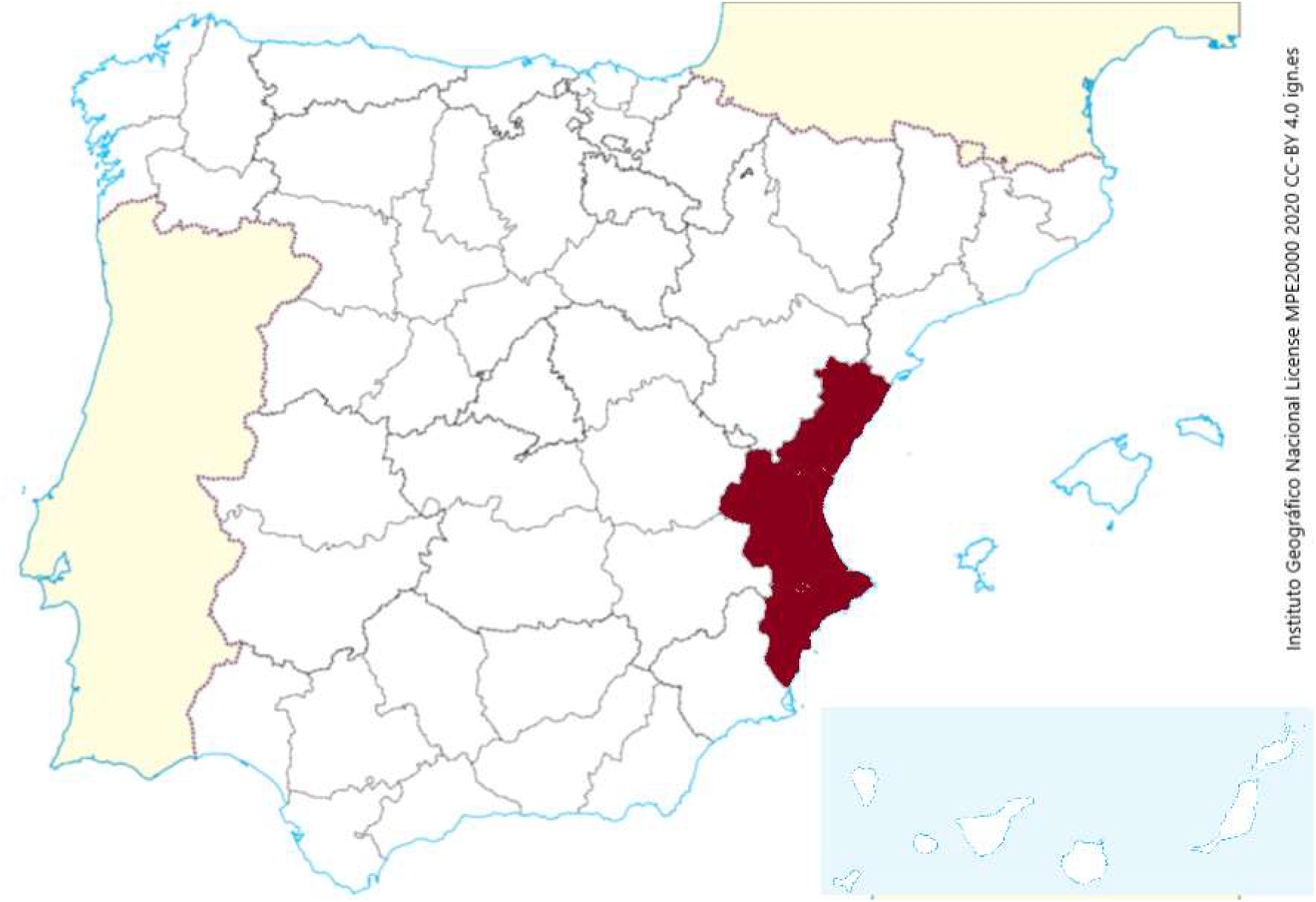
Map of the covered population area in south-east Spain (red). The number of inhabitants covered by the VAHNSI network is 1,118,732.

Samples are tested in one centralized laboratory (FISABIO-Public Health) by real-time reverse-transcription polymerase chain reaction (RT-PCR) for 19 different viruses (influenza A(H1N1)pdm09, A(H3N2), B/Victoria and B/Yamagata, Respiratory Syncytial Virus (RSV) A and B, metapneumovirus, parainfluenza 1-4, rhinovirus/enterovirus, adenovirus, bocavirus, and coronavirus 229E, HKU1, NL63 and OC43). First, all available samples from the 2019/2020 season were re-analysed individually to detect SARS-CoV-2. An in-house assay was rapidly developed in February 2020 based in the WHO-Charité and U.S. CDC assays, to detect the SARS-CoV-2 E-gene (10) in multiplex together with the human cellular control RNAse-P to ensure reliability of sample collection and nucleic acid extraction before RT-PCR. Only samples with RNAse-P *Ct* values <35 were considered valid. The limit of detection of the assay was established to 35 copies using the SARS-CoV-2 E-gene as reference material either as an insert in a pCDNA plasmid (a gift from Luis Enjuanes, Coronavirus laboratory, CNB-CSIC, Madrid, Spain) or as encapsulated RNA (a gift from Charité Medical Center, Berlin, though the European Virus Archive program). Further, to investigate the possibility of SARS-CoV-2 infection earlier to fall-winter 2019 than previously recognized, we retrospectively tested in pools of 10 samples available archived viral RNAs from nasopharyngeal samples collected from 4,010 individuals admitted because of ILI during the 2018/2019 influenza season. The Ethics Research Committee of the Dirección General de Salud Pública-Centro Superior de Investigación en Salud Pública (DGSP-CSISP) previously approved the protocol for the study of respiratory viruses in the VANSHI network.

## Results

A total of 5,176 patients were recruited in the VAHNSI network between November 2019 and mid-March, 2020 and 1,843 were included after applying exclusion criteria; 1,291 of them were PCR-negative for the 19 tested seasonal viruses and the remainder 552 had at least one detectable virus. After re-testing 1,823 available samples for SARS-CoV-2, either positive or negative for seasonal viruses, only one SARS-CoV-2 positive was detected. The result was replicated in an independent sample at the hospital laboratory. Whole-genome sequencing was performed by metagenomic sequencing with spiked primer enrichment, MSSPE (11), using a primer set described by Manning *et al*. (https://www.protocols.io/view/sars-cov-2-enrichment-sequencing-by-spiked-primer-bc36iyre), and showed three amino acid changes from the hCoV-19/Wuhan/WIV04/2019 strain (P323L in NSP12, L117M in NSP8, and D614G in the S protein), belonging to GISAID G clade and PANGOLIN B.1.5 lineage. The infected patient was a man in his late 70’s who was admitted to hospital on early March 2020 (epidemiological week 11/2020) for acute respiratory infection with fever, cough and malaise, with symptom onset 2 days prior to admission. Co-morbidities included diabetes and a prostate adenocarcinoma, but not obesity or smoking history. During hospitalization, he was not admitted in the Intensive Care Unit (ICU) nor required mechanical ventilation or extracorporeal membrane oxygenation (ECMO). However, the patient died five days after admission. As main discharge diagnosis, the ICD10-CM code J06.9 was registered (acute upper respiratory tract infection, unspecified). Finally, all RNA pools from the 4,010 archived samples belonging to the prepandemic 2018/19 influenza season (back to September 2018) were SARS-CoV-2 RT-PCR negative.

## Discussion

To our knowledge, our study is the first comprehensive screening for SARS-CoV-2 in ILI cases at the population level in the prepandemic period in Europe. We describe here the first SARS-CoV-2 admission in the Valencia Region without travel history and thus likely due to community transmission in early March, 2020. The infected patient was an old diabetic man in his late 70’s who was also developing a neoplasia. It is widely known that COVID19 mainly affects elderly people (12, 13) as well as people with underlying chronic conditions (14) including diabetes in an important percentage of severe cases (15, 16). Our patient had a weak health status. Apart from his chronic conditions, he had other frailty indicators (data available upon request). Although the patient was not admitted in the ICU, he finally died 5 days after admission.

The first COVID19 cases in Spain were imported cases detected on 31 January 2020 in a German tourist in La Gomera, Canary Islands and in continental Spain on February 26, in an Italian returning from Milan (3, 4). In the Valencia Region of Spain, the first identified case was also imported (travel history to Nepal in late December 2019), detected in early February 2020 in a Hospital not covered by our surveillance network (17). Excess cases of influenza detected by analysis of primary care electronic medical records suggested an earlier circulation of SARS-CoV-2 in other regions of Spain (8), but no laboratory identification of SARS-CoV-2 was attempted. Retrospective identification of SARS-CoV-2 was also recently suggested in swabs collected for other diagnoses or in hospitalised cases with pneumonia in Spain, so the epidemic may had gone unnoticed prior to first case of local transmission (18). Unfortunately, laboratory data on that observation are yet unpublished.

Countries from the Northern Hemisphere were already engaged in seasonal influenza control when the SARS-CoV-2 outbreak started in China. Therefore, influenza surveillance systems with wide screening windows are perfect tools to retrospectively analyse the presence of the new SARS-CoV-2, because COVID19 shares most of the ILI symptomatology such as fever and cough (12). The first study based in retrospective analysis of samples from ILI cases was performed in Wuhan, China (October 2019-January 2020), detecting the first SARS-CoV-2 cases in the first week of January 2020, concomitant with significantly higher ILI cases for the 2019– 2020 winter in comparison to previous years (19). In contrast, in our network of 4 tertiary-care hospitals in South-eastern Spain (covering a population of more than 1 million), the time of the first non-travel related SARS-CoV-2 hospital admission we detect is early March 2020 and coincided with the end of the peak of influenza and other respiratory viruses (Fig. 2). In addition, ILI inpatient cases for the 2019–2020 winter were not significantly higher in comparison to previous years: average percentage inpatients with ILI were 30% in 2019-20 in our catchment area, as compared to 36% during 2018-19, with no increased ILI cases during the first 10 epidemiological weeks. Phylodynamic analyses estimated the origin of SARS-CoV-2 clusters in Spain around mid-February 2020 (21), which probably later completely replaced circulating seasonal respiratory viruses quickly in our region (Figure 2). In line with our results, SARS-CoV-2 was not detected in similar retrospective studies on inpatients limited to few cases in another region of Spain (n=170; 1 January - 25 February 2020) (20), central Italy (n=166; 1 November 2019 - 1 March 2020) (22), or in only 2/2,888 inpatients and outpatients who had negative results by routine respiratory virus testing in the United States between 1 January 2020 - 26 February 2020 (23). Collectively, our results and published data indicate that the early pandemic phase was still dominated by seasonal respiratory viruses, with very low circulation of SARS-CoV-2 in the community and probably a limited period of overlap with influenza circulation (24-25). Whether pre-existing immune memory and innate immune interference shaped SARS-CoV-2 spread deserves further investigation (24).

**Figure 2.**
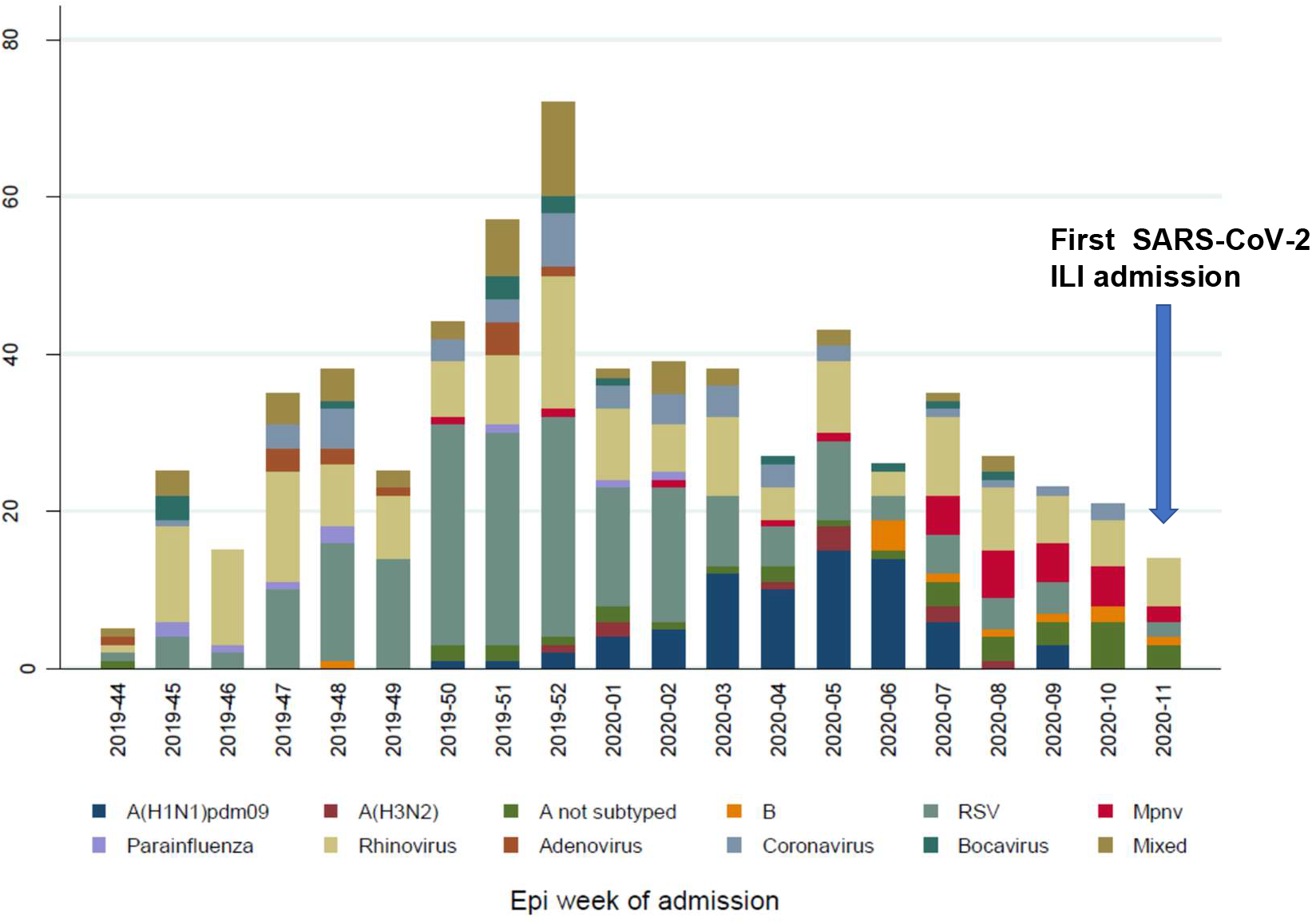
Number of detections for 19 respiratory viruses in the VAHNSI network during the 2019/20 influenza season by epidemiological week (November 1^st^ 2019 to March 14^th^ 2020). The timing for the first ILI admission positive for SARS-CoV-2 is indicated.

The first SARS-CoV-2 cases in hospital admissions in Europe had been identified at the end of January or early February 2020 as related to travel to, or to contacts with, travellers from China (26-28). Later, retrospective analysis identified earlier cases from early or late December 2019 in Italy and France, respectively (29, 30). Some studies have suggested circulation of SARS-CoV-2 may have occurred even earlier than December 2019. A retrospective study on blood samples from Italy found SARS-CoV-2 receptor-binding domain (RBD) specific antibodies as early as September 2019 (30) but whether this observation is due to cross-reactivity with common cold coronaviruses or true SARS-CoV-2 infection is unknown. Previous retrospective sample studies based on RT-PCR testing are limited to 2020 or late December 2019 (22-24, 29, 30). In contrast, we analyzed ILI samples back to September 2018, and found no evidence supporting SARS-CoV-2 circulation in hospitalized community-acquired ILI during the prepandemic 2018/2019 or pandemic 2019/2020 influenza seasons, prior to early March 2020.

Our study has several limitations. First, the VAHNSI network was designed for detecting influenza cases, therefore COVID19 cases presenting a symptomatology different from influenza were not detected. Second, the pandemic disruption situation forced the interruption of hospital surveillance activities in mid-March, just after the declaration of the state of alarm by the Spanish Government. More cases of SARS-CoV-2 would have been detected should the study had not been stopped. Finally, it is possible that pooled testing for 2018-19 samples led to lower RT-PCR sensitivity. However, pool sizes of up to 10 samples have been proven to maintain high performance when compared with individual sample testing (23, 32).

Despite these limitations, here we show how influenza surveillance networks are a powerful resource to analyse the presence of the new respiratory viruses sharing the ILI / SARI symptomatology, because routine collection and storage of samples months (and eventually years) before the occurrence of a new virus introduction. Using these networks will allow us to expand surveillance of SARS-CoV-2, to detect unexpected pathogens in the future, and to elaborate a better map and seasonality for SARS-CoV-2 circulation during the next influenza season. The transition of influenza surveillance networks to other respiratory viruses is thus warranted.

## Data Availability

Data is available under reasonable request

## Acknowledgements

FXLL and BMC is supported by the CIBEResp, Instituto de Salud Carlos III, Spain. Data collection was partially supported by a grant from the Foundation for Influenza Epidemiology (FIE), which supports the Global Influenza Hospital Surveillance Network (see www.gihsn.org for more details). The sponsor did not participate in the study design, data analysis and interpretation, in the writing of the manuscript or in the decision to submit the manuscript for publishing. The authors thank the staff of Hospital General de Castellón, in Castellón; Hospital La Fe, in Valencia; Hospital Doctor Peset, in Valencia; Hospital General de Alicante, in Alicante for their support and contribution to the Valencia Hospital Network for the Study of Influenza and Respiratory Viruses Disease (VAHNSI) network.

## Notes

### Competing Interest Statement

Dr. LOPEZ-LABRADOR reports the work in FISABIO-Public Health was partly funded by Sanofi Pasteur. Sanofi Pasteur did not participate in the design, conduct of the study, analysis or decision to publish the results.
Dr. Mengual-Chulia reports grants from Sanofi Pasteur, during the conduct of the study.
Dr. Mira-Iglesias reports other from Sanofi Pasteur, during the conduct of the study; other from Sanofi Pasteur, outside the submitted work.
Dr. DIEZ-DOMINGO reports grants from SANOFI-PASTEUR, during the conduct of the study; and SP supported teaching activities from my institution.

### Funding Statement

FXLL and BMC are supported by the CIBEResp, Instituto de Salud Carlos III, Spain. Data collection was partially supported by a grant from the Foundation for Influenza Epidemiology (FIE), which supports the Global Influenza Hospital Surveillance Network (see www.gihsn.org for more details). The sponsor did not participate in the study design, data analysis and interpretation, in the writing of the manuscript or in the decision to submit the manuscript for publishing.

### Author Declarations

The Ethics Research Committee of the Direccion General de Salud Publica-Centro Superior de Investigacion en Salud Publica (DGSP-CSISP) previously approved the protocol for the study of respiratory viruses in the VANSHI network.

### Summary of Updates

Typo error corrected in the website abstract on the number of samples analyzed

